# A Probabilistic Approach to Physician-Assisted Suicide

**DOI:** 10.1101/2023.01.24.23284910

**Authors:** Oded Nov

## Abstract

Physician-Assisted Suicide (PAS) is considered by some patients who learn they are at risk for a cognitive decline owing to Alzheimer’s Disease. At the same time, the prospect of PAS may raise patients’ fear of imminent death. Can PAS be offered in a way that is preference-sensitive on one hand, and mitigates patients’ fear of imminent death on the other? A thought experiment of a *Probabilistic Approach to PAS* (Probabilistic PAS) is proposed here as a possible solution in jurisdictions where PAS is legal.

Consider the following scenario: A patient diagnosed with Alzheimer’s Disease who is considering PAS might request that when he or she can no longer recognize their loved ones, their doctor will give them a lethal pill that has a 1/100 daily probability of being activated. As a result, after taking the pill, every day the patient will have a 1/100 probability of dying. The likelihood of the patient dying within a year from taking the pill is 97.4%, and within two years, it is 99.9%. As such, a Probabilistic PAS with which on any given day the probability of dying is low, can help patients avoid the fear of imminent death – which traditional PAS entails – while respecting their preference to end their lives.

To examine the potential reception of a Probabilistic PAS, a survey was administered to a nationally-representative sample of US residents, using Prolific, a research participant recruitment platform. 499 participants were presented with a short description of a patient who was diagnosed with Alzheimer’s Disease, is writing an advance directive and is considering ways to end their life painlessly when they can no longer recognize their loved ones. Participants were asked about their own preferences in case they were to face a similar situation, and whether helping administer Probabilistic PAS would be ethical for the patient’s provider.

498 participants responded to the question about their own preference. 73.5% indicated that they would choose one of the two PAS options. Among those, 9.8% preferred a Probabilistic PAS over traditional PAS. Men were more likely than women to favor Probabilistic PAS for themselves. 482 participants indicated which option would be most ethical for the patient’s provider to administer. 48.1% indicated one of the two PAS options as most ethical. Among those, 10.3% considered Probabilistic PAS to be more ethical than traditional PAS. Men were more likely than women to consider the provider administering Probabilistic PAS to be most ethical.

A version of the Probabilistic PAS proposed here should be considered as a preference-sensitive option presented by healthcare providers to patients considering advance care planning in places where PAS is available.

## Introduction

Physician-Assisted Suicide (PAS) is considered by some patients who learn they are at risk for a cognitive decline owing to Alzheimer’s Disease^1,2^. At the same time, the prospect of PAS may raise patients’ fear of imminent death. Can PAS be offered in a way that is preference-sensitive on one hand, and mitigates patients’ fear of imminent death on the other? A thought experiment of a *Probabilistic Approach to PAS* (Probabilistic PAS) is proposed here as a possible solution in jurisdictions where PAS is legal, and the results of a US nationally representative sample survey of responses to it are presented.

## Methods

To examine the potential reception of a Probabilistic PAS, a survey was administered to a nationally-representative sample of US residents, using Prolific, a research participant recruitment platform. 499 participants (252 women; 240 men; 7 other; mean age: 46.2, SD: 16.6) were presented with a short description of a patient who was diagnosed with Alzheimer’s Disease, is writing an advance directive and is considering ways to end their life painlessly when they can no longer recognize their loved ones^3^. Participants were asked about their own preferences in case they were to face a similar situation, and whether helping administer Probabilistic PAS would be ethical for the patient’s provider. In responding to the questions, participants had to choose between three alternatives including traditional PAS, a Probabilistic PAS, and no PAS. The choice order presented to participants was randomized. The study was approved by NYU Institutional Review Board (IRB-FY2022-6529).

## Results

498 participants (252 women; 239 men; 7 other) responded to the question about their own preference. 366 (73.5%) indicated that they would choose one of the two PAS options (traditional PAS and Probabilistic PAS). Among those, 36 (9.8%) preferred a Probabilistic PAS over traditional PAS. Men were more likely than women to favor Probabilistic PAS for themselves (23/239 vs. 13/252; p<0.01). 482 participants (242 women; 233 men; 7 other) indicated which option would be most ethical for the patient’s provider to administer. 232 (48.1%) indicated one of the two PAS options as most ethical. Among those, 24 (10.3%) considered Probabilistic PAS to be more ethical than traditional PAS. Men were more likely than women to consider the provider administering Probabilistic PAS to be most ethical (18/233 vs. 6/241; p<0.01). In both cases, age was negatively associated with a PAS preference over no PAS (p<0.01). Age was not associated with a Probabilistic PAS preference.

## Discussion

A Probabilistic PAS is outlined as a thought experiment to inform the scholarly discussion of PAS^4,5^. Probabilistic PAS represents an alternative to the life/death binary choice currently available to patients in jurisdictions were PAS is legal, and offers an additional avenue for decision making autonomy. To examine its possible reception, Probabilistic PAS was presented to a US nationally representative sample. The Probabilistic PAS option was preferred by a minority of those who indicated a preference for PAS if they were to learn they are likely to suffer from Alzheimer’s Disease. A version of the Probabilistic PAS proposed here should be considered as a preference-sensitive option presented by healthcare providers to patients considering advance care planning in places where PAS is available^6^.

## Data Availability

All data produced in the present study are available upon reasonable request to the authors

